# When Data Reform Meets Bureaucratic Hierarchy: A Political Economy Analysis of Institutionalizing a District Health Data Bank in West Sumbawa District, Indonesia

**DOI:** 10.64898/2026.08.25.26361308

**Authors:** Muhammad Asrullah, Abigael Wohing Ati, Dhea Keyle Fortunandha, Garin Frige Janitra, Ery Setiawan, Ulfathea Mulyadita, Astri Pratiwi, Shita Listya Dewi, Matt Boxshall

**Affiliations:** Centre for Health Policy and Management, Faculty of Medicine, Public Health, and Nursing, Universitas Gadjah Mada, Indonesia; Division of Human Nutrition and Health, Wageningen University and Research, the Netherlands; Health Systems Insight, Indonesia; Faculty of Public Health, Universitas Indonesia, Indonesia; Health Systems Insight, Washington DC, USA

**Author notes:** Corresponding author* Muhammad Asrullah. These authors contributed equally to this work and share first authorship (Muhammad Asrullah, Abigael Wohing Ati).

**Keywords:** Health Data Bank, Data Institutionalization, Political Economy, District Health Governance, Indonesia

## Abstract

**Background:** Although often perceived as a technical tool, a data bank is fundamental for district performance management, facilitating integrated coordination, data consolidation, and routine information use for decision-making. However, implementation unfolds within hierarchical bureaucratic systems shaping authority distribution, workload allocation, coordination, and resource use. This study examines the political economy of institutionalizing a district-level health data bank in West Sumbawa District, Indonesia.

**Method:** A longitudinal qualitative case study was conducted in West Sumbawa District, West Nusa Tenggara Province, Indonesia, from November 2025 to March 2026 across three evaluation phases (baseline, midline, endline), involving 25 District Health Office (DHO) officers appointed to the Health Data Bank team, representing five organizational units, including the Secretariat, General and Human Resources Unit, and Public Health Division. Data were collected through participatory workshops, focus group discussions, in-depth interviews, observation, and document review, including official decrees, SOPs, meeting minutes, and implementation records, and analysed using Bossert’s Decision Space Framework combined with a problem-driven political economy analysis examining how power relations, institutional norms, workload, resource support, and perceived incentives influenced whether technical reforms became operationalized in routine practice.

**Results:** The Health Data Bank progressed from strong institutional acceptance to structural formalization. Decision-making authority remained centralized within the Secretariat, enabling coordination but limiting distributed ownership. Resource constraints increased workload, concentrated in the Secretariat and division coordinators responsible for data consolidation and validation, without dedicated financing or staffing, despite improved analytical capacity. Accountability mechanisms were established through governance instruments, though enforcement and feedback loops remained underdeveloped. Data submission, validation, and use were not yet fully institutionalized, resulting in a gap between structural readiness and functional use.

**Conclusion:** Data institutionalization involves both technical and organizational processes, requiring collaborative negotiation of authority, workload, and resources. Continued attention to these factors will help structural formalization translate into sustainable operational outcomes.

## Introduction

Health data bank is fundamental in district performance management reforms, but frequently conceptualized through technical approaches to data integration, emphasizing interoperability, digital platforms, and standardized reporting. When implemented within a bureaucratic systems, however, the outcomes are shaped not only by technical design but also by institutional and political-economic dynamics.

A health data bank, as used in this study, is an integrated data platform that consolidates indicators from multiple programs into a single system. It links existing digital data sources, gives staff a shared point to validate and review records, and presents results through dashboards for planning and monitoring. It is not simply a storage device. Building and validating the initial links between program datasets requires substantial staff time for data processing, entry, and analysis; the intended payoff is that routine use later reduces the manual compilation work described below. In West Sumbawa District, the DHO developed the Health Data Bank, locally branded MATA SIDIK, specifically for its own context as part of a broader performance management strengthening program; it is not a commercial off-the-shelf product. The sections that follow describe what data the platform links, who inputs and uses it, and the additional workload this process created for district staff.

In decentralized health systems across low- and middle-income countries (LMICs), District Health Offices (DHOs) occupy a critical intermediary role. Positioned closest to service delivery, they are responsible for translating national health priorities into local action while simultaneously producing performance data for higher-level decision-making. Despite this dual mandate, DHOs often operate within fragmented reporting and data management arrangements, where multiple vertical reporting structures coexist with limited coordination [1–3].

Over the past two decades, substantial investments have been directed toward integrated digital health and data management platforms, such as the District Health Information System 2 (DHIS2) [4]. These efforts are grounded in the assumption that improved technological infrastructure is necessary, though not sufficient, for enhanced data use. However, empirical evidence suggests that technical integration alone does not guarantee data-driven decision-making. Even where systems are functional, data use at the district level remains inconsistent and shaped by organizational factors beyond system design [5–7].

Emerging scholarship has reframed this implementation gap through a political economy lens. This perspective moves beyond technical capacity to examine how authority structures, resource allocation, workload distribution, and institutional incentives influence reform trajectories [8,9]. In hierarchical bureaucracies, reforms are embedded within existing power relations and administrative routines, which shape how new approaches are adopted, coordinated, and sustained over time.

Indonesia provides a particularly relevant context for examining these dynamics. Following decentralization reforms, DHOs hold formal authority over planning, monitoring, and service coordination. However, this authority is constrained by vertically organized reporting systems, overlapping staff responsibilities, and limited flexibility in budget allocation. National reforms such as the Satu Data Indonesia initiative aim to promote integrated data governance, yet translating these frameworks into routine operational practice at the district level remains challenging [10,11].

In parallel, the Government of Indonesia has introduced the Integrated Primary Health Care (ILP) approach as part of broader health system transformation efforts [12]. ILP aims to shift service delivery from fragmented, program-based approaches toward a more integrated, life-cycle-based model of care, strengthening continuity of services across promotive, preventive, curative, and rehabilitative functions at the primary care level. This includes efforts to improve how districts consolidate, review, interpret, and use data across programs to support planning, monitoring, and adaptive decision-making. However, districts often face challenges in obtaining a consolidated and actionable view of health system performance because data remain dispersed across multiple program-specific reports, databases, and reporting channels. As a result, managers frequently rely on time-consuming manual compilation processes to generate information for planning, monitoring, and performance review. To address these challenges, the Health Data Bank was introduced in West Sumbawa District as part of a broader district performance management strengthening approach. The initiative was designed to serve as a single platform for consolidating, validating, reviewing, and using priority indicators across programs, thereby supporting integrated data governance and routine performance management processes. The initiative was not a standalone digital system; it aimed to strengthen evidence-informed decision-making within routine district management processes.

Against this backdrop, the implementation process in West Sumbawa District provides an opportunity to examine how organizational and political-economic dynamics shape the institutionalization of district-level performance management reforms. While the initiative received strong leadership endorsement and demonstrated growing institutional acceptance, implementation also revealed challenges related to authority distribution, workload concentration, coordination, and sustainability.

This paper examines how political economy factors, including power relations, institutional support, and perceived benefits, shape the institutionalization trajectory of a district-level Health Data Bank initiative as a part of District Performance Management reform. Drawing on Bossert’s Decision Space Framework and a problem-driven political economy perspective, the study analyzes how authority, resources, accountability, and organizational structures influenced implementation across three longitudinal phases. We argue that institutionalizing district performance management is not merely a technical process, but an organizational negotiation shaped by governance arrangements and operational realities.

## Methods

### Study design

This study applies a longitudinal qualitative case study design to examine the political economy of institutionalizing a district-level health data bank in Indonesia. Data were drawn from a series of evaluation studies conducted between November 2025 and March 2026 across three implementation phases: Phase 1 (Preparation and Development, December 2025), Phase 2 (Operation and Early Implementation, January 2026), and Phase 3 (Results and Utilization, February--March 2026). The evaluation was guided by a political economy lens, drawing on Bossert’s decision space framework to examine how authority distribution, workload allocation, resource availability, and institutional incentives shaped the implementation trajectory. This analysis was conducted to surface organizational and structural dynamics that a technical implementation framework alone cannot fully capture.

To strengthen rigor and credibility, triangulation was conducted across multiple data sources and methods, including participatory workshops, focus group discussions, in-depth interviews, observations, and document reviews [13]. Information obtained from one source or method was compared and cross-checked with findings from other methods and respondents across different implementation phases. For example, findings emerging from interviews were verified through workshop discussions, implementation observations, meeting minutes, and official documents such as SOPs and decrees. Codes and categories generated by one researcher were checked, discussed, and confirmed by the others to strengthen analytical consistency and interpretation.

### Study Setting

The study was conducted at the DHO of West Sumbawa District, West Nusa Tenggara Province, Indonesia. The Health Data Bank initiative was introduced here as part of a broader health systems strengthening program supported by Health System Insight Indonesia (HSI), with technical and evaluative support from the Centre for Health Policy and Management, Universitas Gadjah Mada (CHPM UGM).

Participants were drawn from the formally established Health Data Bank Team, comprising 25 staff members appointed through an official decree (*Surat Keputusan*, SK). The team represented five organizational units within the DHO: the Secretariat, the General and Human Resources Unit, the Public Health Division (*Kesmas*), the Disease

Prevention and Control Division (P3KL), and the Health Services and Resources Division (PMSDK). All team members participated across at least one evaluation phase, with the Head of DHO and senior division coordinators participating across multiple phases.

### Conceptual Framework

This study applies an integrated analytical framework combining a Bossert’s Decision Space Framework with a problem-driven political economy lens focusing on how authority structures, workload distribution, resource allocation, institutional incentives, and coordination dynamics shape the institutionalization of a district-level health data system [14,15]. Bossert’s framework conceptualizes decentralization as the range of choice (decision space) exercised by local actors across key dimensions of authority, institutional capacity, and accountability [16]. Bossert distinguishes de jure decision space, the authority formally assigned by higher levels of government, from de facto decision space, the authority actually exercised in practice [17]. In this study, these dimensions are operationalized through four system domains: data and information systems, human resource capacity and data analysis, performance management ecosystem, and accountability and governance. Data systems and performance ecosystems primarily reflect authority over coordination and decision-making processes; human resource capacity represents the institutional capacity dimension of Bossert’s framework, specifically the staffing and time available to exercise decision space; and accountability and governance correspond to oversight and enforcement mechanisms.

Building on Bossert’s framework, this study applies a political economy perspective to explain how formal decision space was shaped in practice by bureaucratic hierarchy, institutional norms, incentive structures, and resource constraints. While decision space identifies the formal range of authority available to local actors, political economy analysis helps explain why this authority was unevenly exercised, operationalized, or sustained during implementation.

Politics and political economy play a central role in shaping whether and how reforms are implemented in both developing and developed countries [14]. Bossert explains where decision space exists, while political economy explains why actors are or are not able/willing to exercise it. Bureaucratic hierarchy, operating as an institutional norm, regulates how authority is concentrated or distributed, and power dynamics within that hierarchy make its effects more complicated. Workload, defined here as the additional time staff spend on data consolidation, validation, communication, and analysis beyond their routine duties, and access to resources reflect institutional support. Perceived incentives, including individual and organizational benefits, importance, and relevance, are foundational to accountability mechanisms and compliance behaviours. The framework further argues that reform outcomes depend on the alignment between power, institutional support, norms and regulations, as well as perceived benefits, with the corresponding fiscal, human resources quantity and capacity, and managerial autonomy. When authority remains centralized while responsibilities are diffused without adequate resource allocation, implementation may achieve formal compliance but stall before reaching functional institutionalization (Fig 1). This framework enables analysis not only of what institutional structures are established, but also of how and why they are (or are not) translated into routine practice within complex bureaucratic systems.

**Fig 1.**
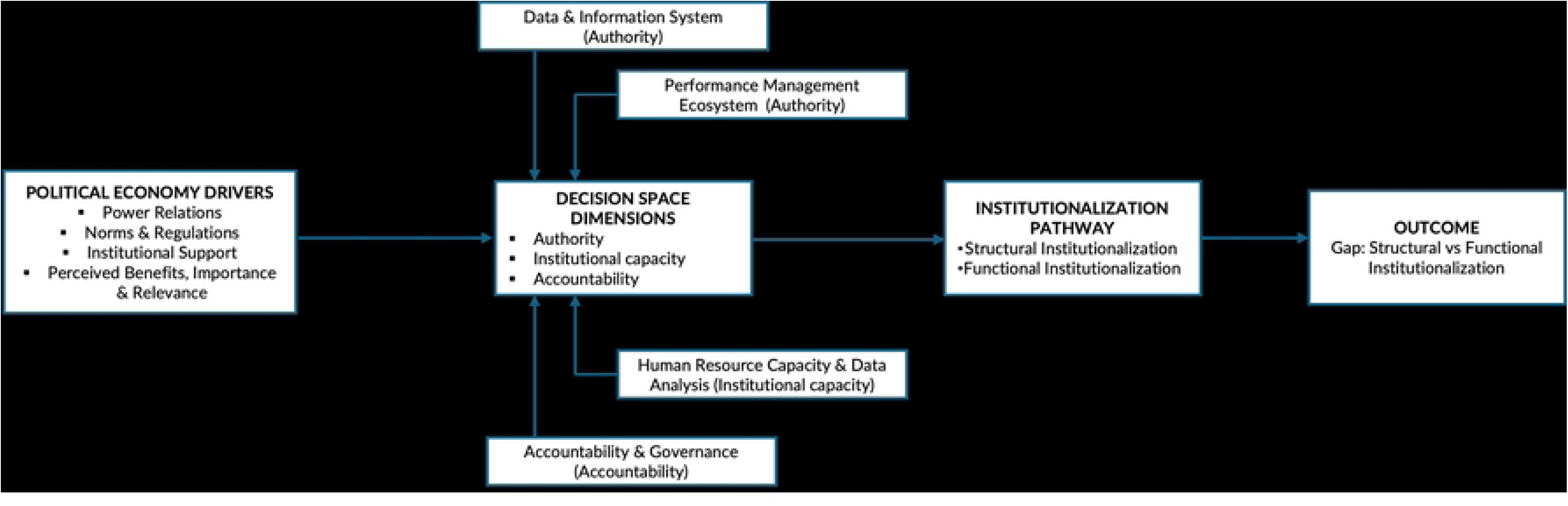
Integrated conceptual framework combining political economy and Bossert’s decision space. Institutionalization unfolds in two stages. Structural institutionalization refers to the establishment of formal elements, such as a decree (SK), SOPs, designated teams, and capacity-building, which signal strong organizational commitment and readiness. However, these structures do not automatically translate into practice. Functional institutionalization occurs only when these formal arrangements are activated through routine processes, including regular data submission, validation, and use in decision-making.

### Data Collection

Data were collected using a combination of methods calibrated to the focus of each phase. In Phase 1, a two-day participatory workshop and key informant interviews (KIIs) were conducted with DHO division representatives, capturing early institutional perceptions, expectations, and the process of indicator and SOP development. Puskesmas staff were included at this early stage to capture how frontline facilities interact with data later consolidated in the Health Data Bank; the formally appointed 25-member Health Data Bank team, and the results reported below, are based at the DHO level. In Phase 2, focus group discussions (FGDs) and structured group discussions explored operational readiness, coordination challenges, and emerging workload dynamics across divisions. In Phase 3, in-depth interviews (IDIs) were conducted with division coordinators, Secretariat representatives, and DHO leadership, focusing on implementation status, authority structures, and perceived sustainability. Document review complemented all phases, encompassing the official decree, SOPs, meeting minutes, dashboard templates, and budget and planning records. A summary of qualitative data collection activities across phases is presented in Table 1.

**Table 1.**
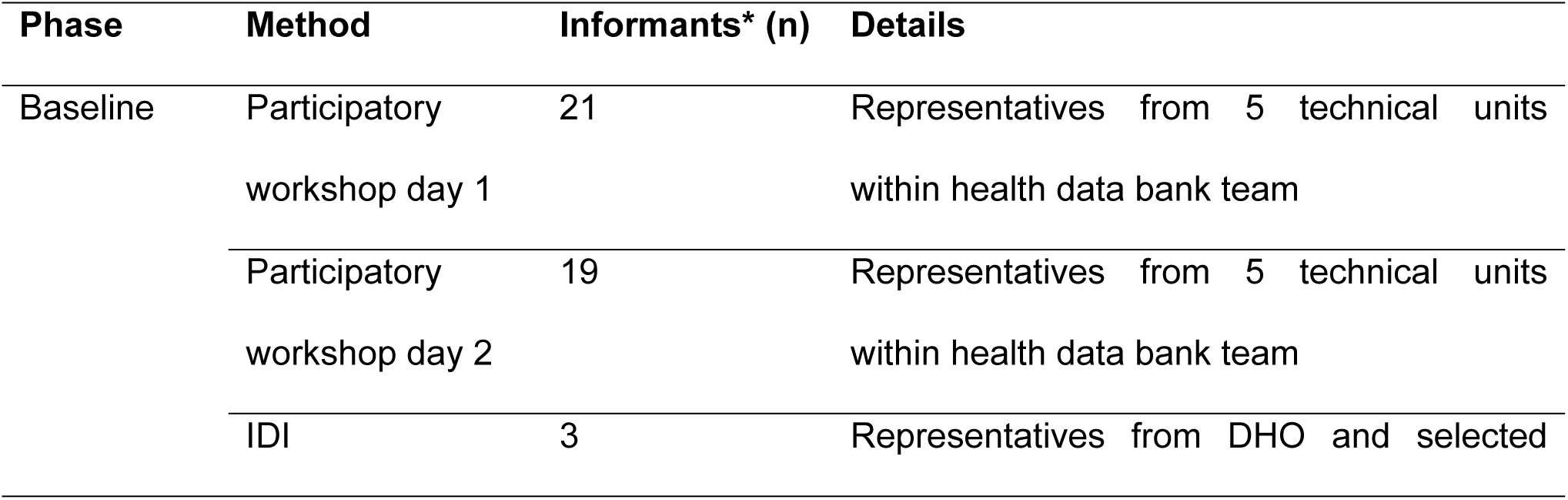
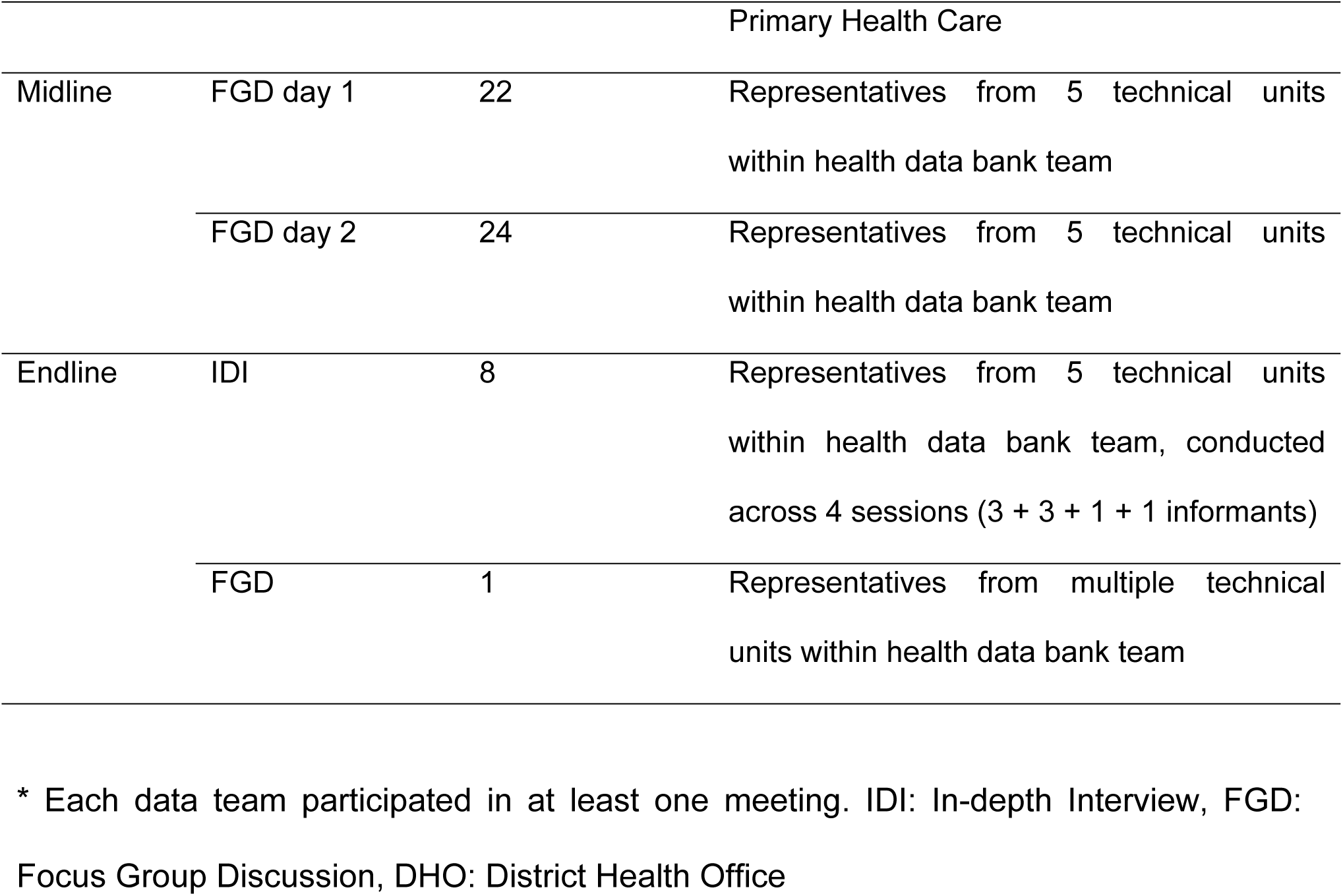
List of qualitative data collection activities.

Data collection tools (interview guides and discussion protocols) were developed based on the study objectives and existing implementation frameworks, and were iteratively refined during early field engagement. Initial tools were pilot-tested during the first phase of data collection (workshops and KIIs), allowing the research team to adjust question clarity, sequencing, and relevance to the local context and phase. While the study applied a longitudinal qualitative design, data collection methods were intentionally adapted across phases to capture evolving implementation processes and organizational dynamics. This approach is consistent with qualitative and implementation research, where multiple methods and triangulation are used to strengthen contextual understanding and analytical rigor across different stages of implementation [13]. The study did not seek strict procedural uniformity. It maintained conceptual continuity by consistently examining the same core analytical domains across phases, including authority, coordination, workload, accountability, and routine data use.

### Data Analysis

All qualitative data, including transcripts, field notes, workshop outputs, and document review, were analysed using a thematic approach following Braun and Clarke’s six-step process of familiarization, coding, theme development, theme review, theme definition, and interpretation [18]. Coding followed a combined deductive and inductive strategy: deductive codes corresponded to the six Proctor domains used in the original evaluation, while inductive codes captured emergent themes related to authority concentration, workload burden, incentive structures, and governance formalization. A traceability matrix was used throughout to systematically link analytical questions, data sources, illustrative quotations, and interpretive conclusions, ensuring transparency and rigor across phases.

All researchers had prior training and experience in qualitative research and were familiarized with the study framework and codebook prior to analysis. A preliminary codebook was developed deductively and refined inductively during early coding. Discussions were held regularly throughout the process to maintain alignment and consistency across coders. Discrepancies were discussed and resolved through consensus, leading to iterative refinement of code definitions.

For this paper, coded material was subsequently interpreted through Bossert’s decision space framework, examining how the distribution of authority, resources, and accountability across organizational levels shaped the pace and depth of institutionalization. Cross-phase comparison was used to identify longitudinal shifts in institutional dynamics and to distinguish between structural formalization and functional operationalization, a distinction central to the analysis.

### Ethical Considerations

The study received ethics approval from the Internal Review Board of the Faculty of Medicine, Public Health, and Nursing at Universitas Gadjah Mada, Indonesia (Approval Number: KE/FK/1275/EC 2025). All participants were informed of the evaluation purpose and provided consent for their contributions to be used for research and learning purposes. Data were anonymized prior to analysis and are reported without personally identifying information.

## Results

A total of 25 participants were included across the three implementation phases, representing key organizational levels within the District Health Office, including leadership, Secretariat, and program divisions. The composition of respondents reflects the institutional structure through which the Health Data Bank was implemented, capturing perspectives from actors with varying roles in decision-making, data management, and program coordination. Table 2 summarizes the characteristics of study participants.

**Table 2.** Characteristic of Informants.

| Characteristics | n (25) |
| --- | --- |
| Sex |  |
| Female | 16 |
| Male | 9 |
| Position |  |
| Team leader (head of DHO) | 1 |
| Secretary | 1 |
| Division coordinator | 5 |
| Division member | 18 |
| Division |  |
| Secretariat | 7 |
| General and Human Resources Unit | 1 |
| Public Health Division (*Kesmas*) | 5 |
| Disease Prevention and Control Division (P3KL) | 6 |
| Health Services and Resources Division (PMSDK). | 6 |

Our study found, across the three implementation phases, decision-making authority over how the Health Data Bank itself was designed, sequenced, and operationalized, not decisions about health programs informed by the data it produces, remained largely concentrated at the leadership level, which facilitated early endorsement and coordination of the Health Data Bank. At the same time, the distribution of authority to division-level actors was gradually developing. In terms of resources, implementation progressed alongside increasing demands on existing staff, with capacity-building activities completed and technical readiness established.

Accountability mechanisms for overseeing implementation of the Health Data Bank itself were established through governance instruments and monitoring processes. Oversight of downstream health service performance informed by Health Data Bank outputs falls outside the scope of this study. While these mechanisms were in place, routine feedback loops and data-informed decision-making processes were still emerging.

A summary of these findings is presented in Table 3. The table presents the intersection between Bossert’s governance dimensions and PEA factors. Bossert’s dimensions identify key areas of decision-making authority, accountability, and resource allocation, while the PEA factors explain the political, institutional, and behavioural dynamics that influence how these decision spaces are exercised and experienced by different actors.

**Table 3.**
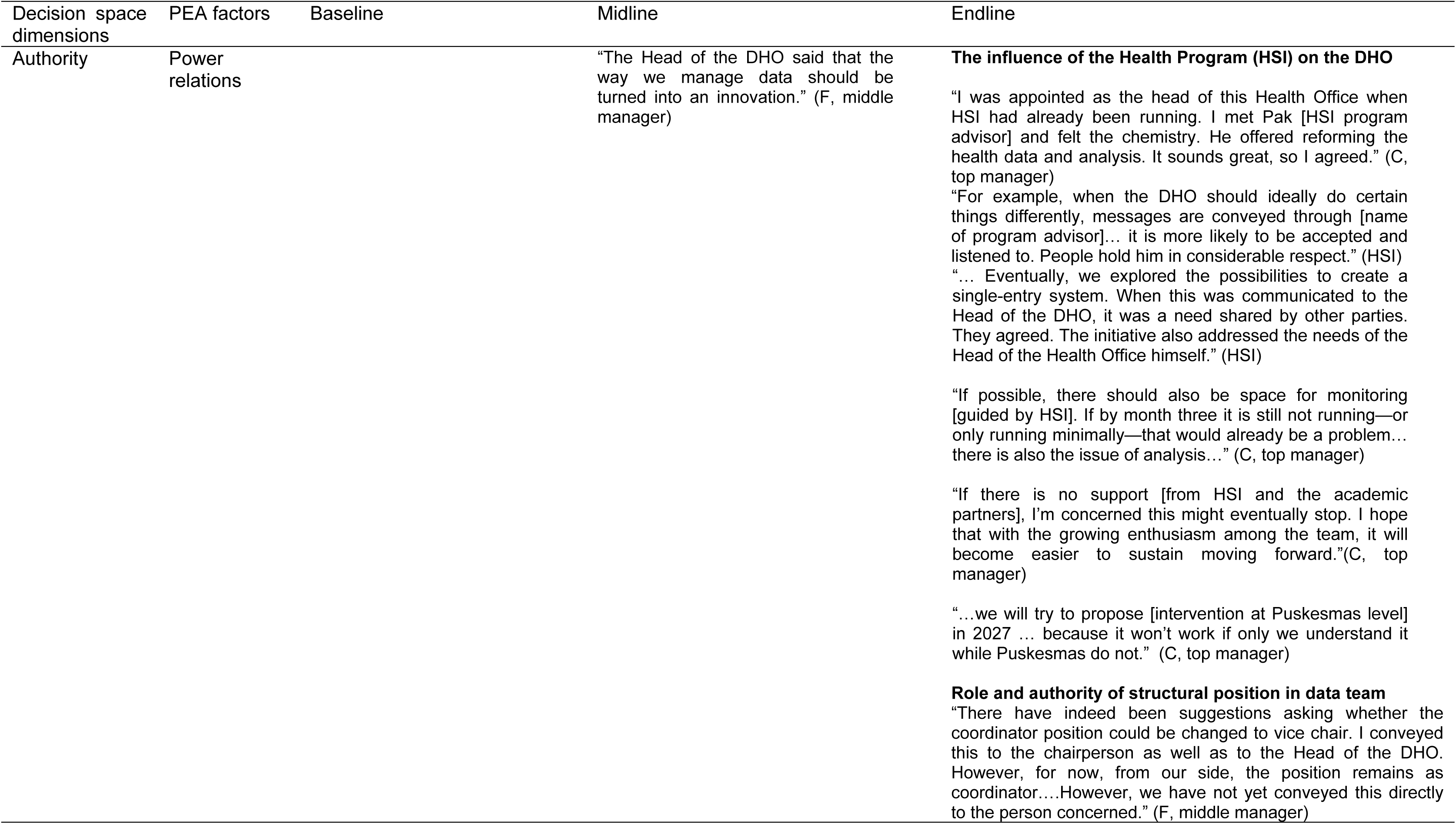
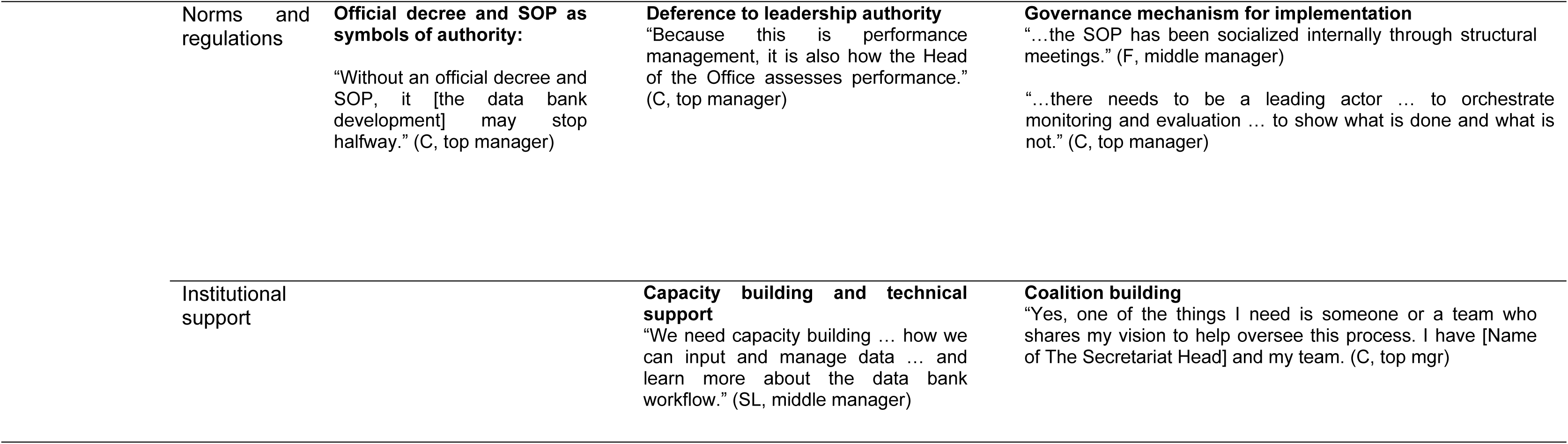
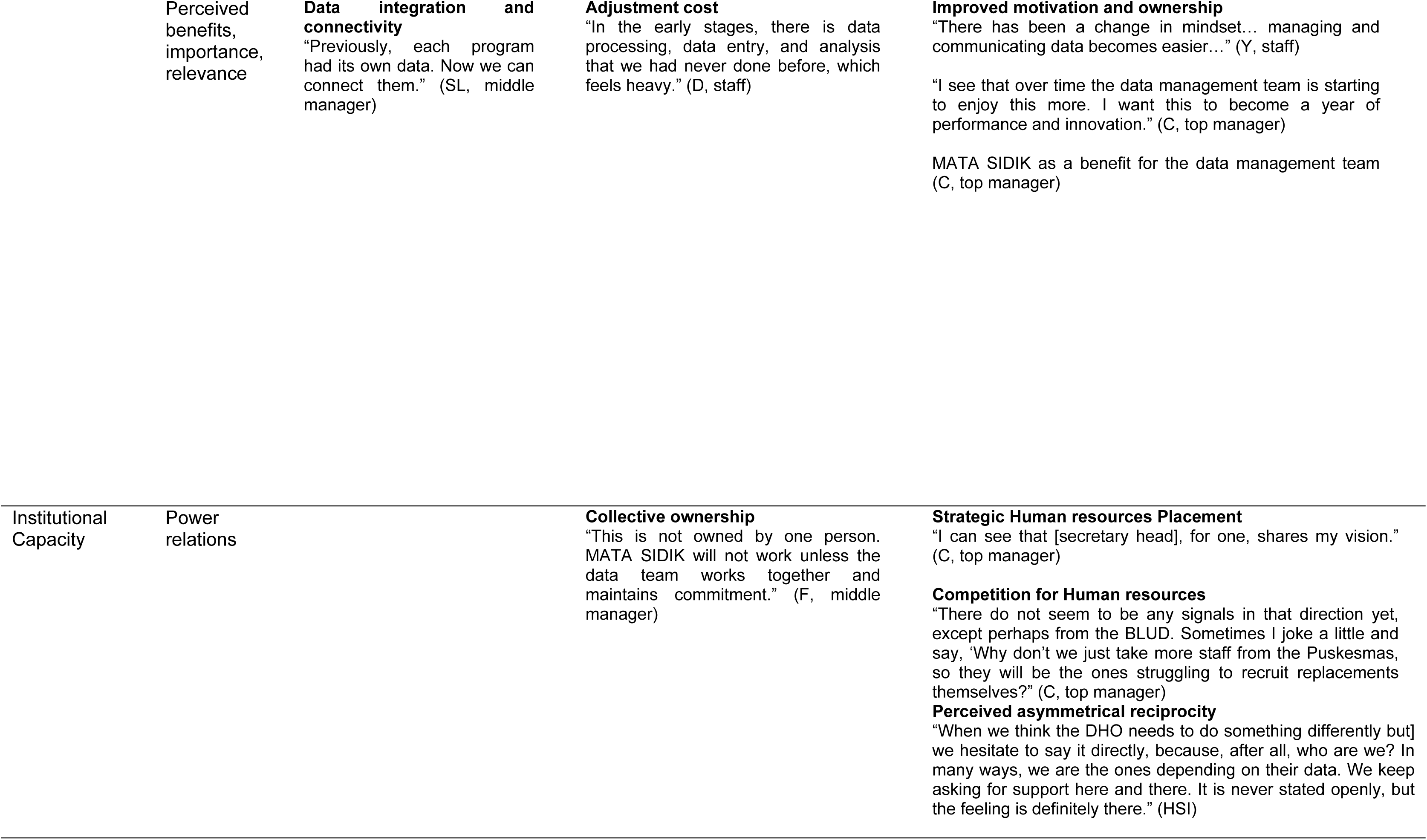
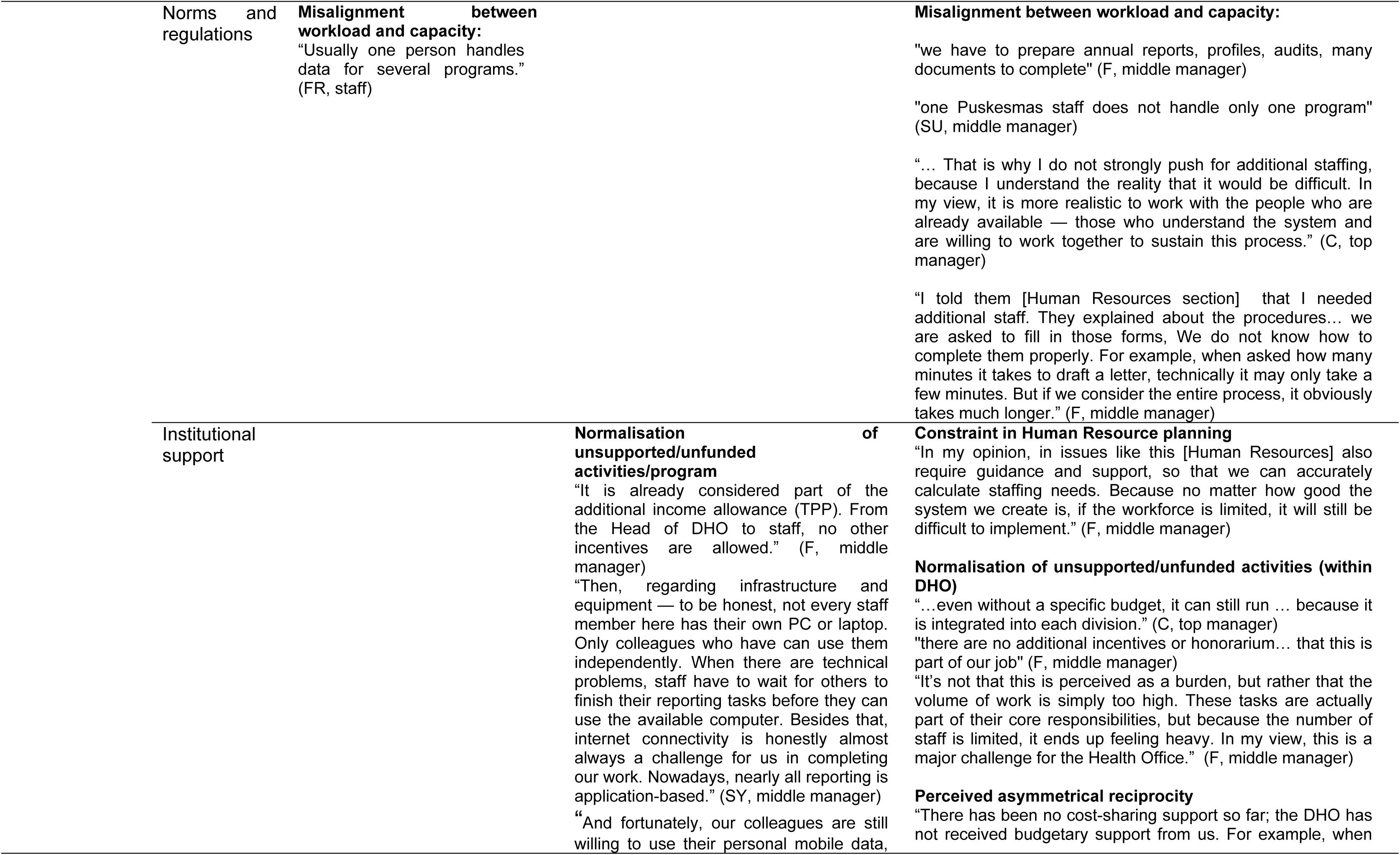
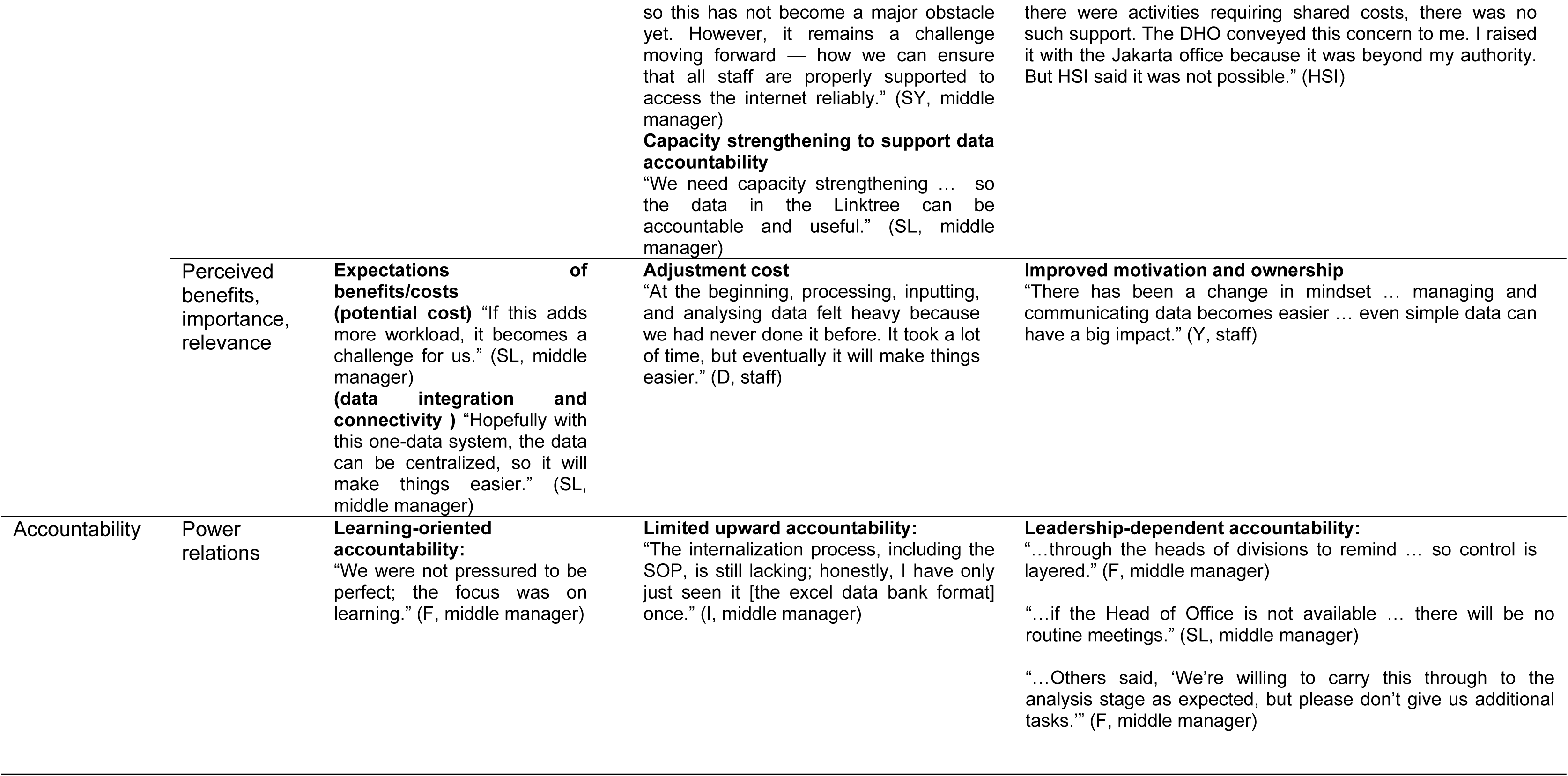
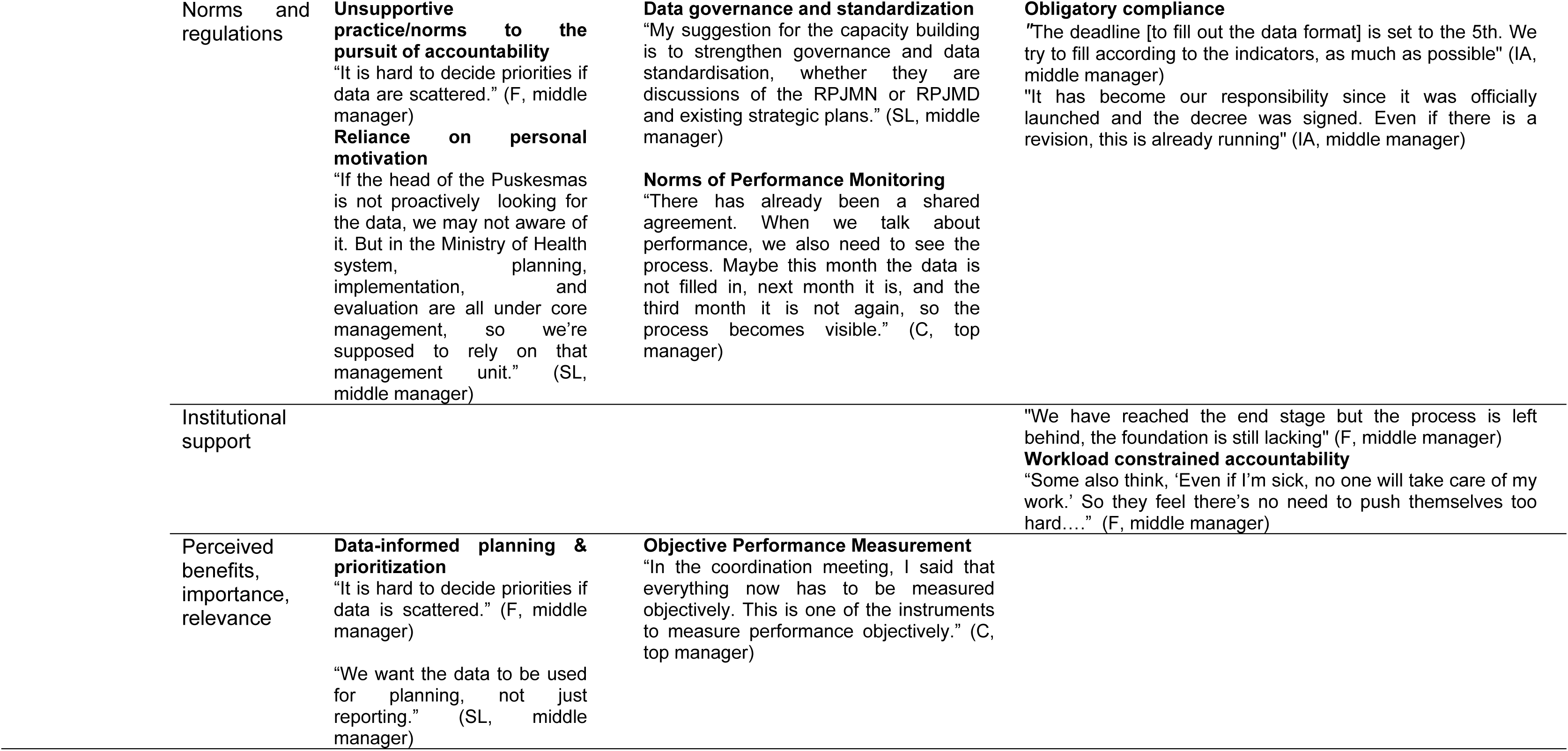
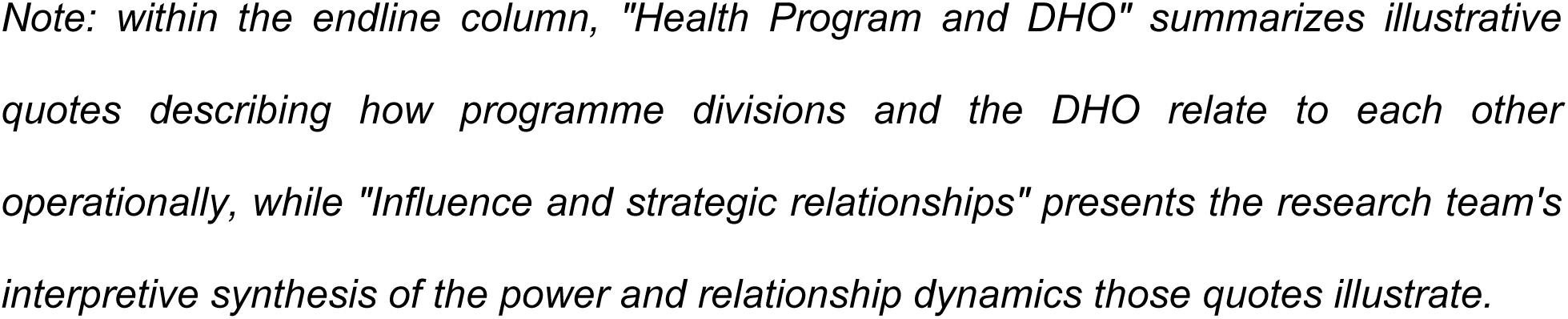
Summary of Findings Organized by Decision Space Domains (Authority, Institutional Capacity, and Accountability)

| Decision space dimensions | PEA factors | Baseline | Midline | Endline |
| --- | --- | --- | --- | --- |
| Authority | Power relations |  | "The Head of the DHO said that the way we manage data should be turned into an innovation." (F, middle manager) | **The influence of the Health Program (HSI) on the DHO**<br>"I was appointed as the head of this Health Office when HSI had already been running. I met Pak [HSI program advisor] and felt the chemistry. He offered reforming the health data and analysis. It sounds great, so I agreed." (C, top manager)<br>"For example, when the DHO should ideally do certain things differently, messages are conveyed through [name of program advisor]... it is more likely to be accepted and listened to. People hold him in considerable respect." (HSI)<br>"... Eventually, we explored the possibilities to create a single-entry system. When this was communicated to the Head of the DHO, it was a need shared by other parties. They agreed. The initiative also addressed the needs of the Head of the Health Office himself." (HSI)<br>"If possible, there should also be space for monitoring [guided by HSI]. If by month three it is still not running—or only running minimally—that would already be a problem... there is also the issue of analysis..." (C, top manager)<br>"If there is no support [from HSI and the academic partners], I'm concerned this might eventually stop. I hope that with the growing enthusiasm among the team, it will become easier to sustain moving forward." (C, top manager)<br>"...we will try to propose [intervention at Puskesmas level] in 2027 ... because it won't work if only we understand it while Puskesmas do not." (C, top manager)<br>**Role and authority of structural position in data team**<br>"There have indeed been suggestions asking whether the coordinator position could be changed to vice chair. I conveyed this to the chairperson as well as to the Head of the DHO. However, for now, from our side, the position remains as coordinator....However, we have not yet conveyed this directly to the person concerned." (F, middle manager) |
|  | Norms and regulations | **Official decree and SOP as symbols of authority:**<br>“Without an official decree and SOP, it [the data bank development] may stop halfway.” (C, top manager) | **Deference to leadership authority**<br>“Because this is performance management, it is also how the Head of the Office assesses performance.” (C, top manager) | **Governance mechanism for implementation**<br>“...the SOP has been socialized internally through structural meetings.” (F, middle manager)<br>“...there needs to be a leading actor ... to orchestrate monitoring and evaluation ... to show what is done and what is not.” (C, top manager) |
|  | Institutional support |  | **Capacity building and technical support**<br>“We need capacity building ... how we can input and manage data ... and learn more about the data bank workflow.” (SL, middle manager) | **Coalition building**<br>“Yes, one of the things I need is someone or a team who shares my vision to help oversee this process. I have [Name of The Secretariat Head] and my team. (C, top mgr) |
|  | Perceived benefits, importance, relevance | **Data integration and connectivity**<br>“Previously, each program had its own data. Now we can connect them.” (SL, middle manager) | **Adjustment cost**<br>“In the early stages, there is data processing, data entry, and analysis that we had never done before, which feels heavy.” (D, staff) | **Improved motivation and ownership**<br>“There has been a change in mindset... managing and communicating data becomes easier...” (Y, staff)<br>“I see that over time the data management team is starting to enjoy this more. I want this to become a year of performance and innovation.” (C, top manager)<br>MATA SIDIK as a benefit for the data management team (C, top manager) |
| Institutional Capacity | Power relations |  | **Collective ownership**<br>“This is not owned by one person. MATA SIDIK will not work unless the data team works together and maintains commitment.” (F, middle manager) | **Strategic Human resources Placement**<br>“I can see that [secretary head], for one, shares my vision.” (C, top manager)<br>**Competition for Human resources**<br>“There do not seem to be any signals in that direction yet, except perhaps from the BLUD. Sometimes I joke a little and say, ‘Why don’t we just take more staff from the Puskesmas, so they will be the ones struggling to recruit replacements themselves?’” (C, top manager)<br>**Perceived asymmetrical reciprocity**<br>“When we think the DHO needs to do something differently but] we hesitate to say it directly, because, after all, who are we? In many ways, we are the ones depending on their data. We keep asking for support here and there. It is never stated openly, but the feeling is definitely there.” (HSI) |
|  | Norms and regulations | **Misalignment between workload and capacity:**<br>"Usually one person handles data for several programs."<br>(FR, staff) |  | **Misalignment between workload and capacity:**<br>"we have to prepare annual reports, profiles, audits, many documents to complete" (F, middle manager)<br>"one Puskesmas staff does not handle only one program" (SU, middle manager)<br>"... That is why I do not strongly push for additional staffing, because I understand the reality that it would be difficult. In my view, it is more realistic to work with the people who are already available — those who understand the system and are willing to work together to sustain this process." (C, top manager)<br>"I told them [Human Resources section] that I needed additional staff. They explained about the procedures... we are asked to fill in those forms, We do not know how to complete them properly. For example, when asked how many minutes it takes to draft a letter, technically it may only take a few minutes. But if we consider the entire process, it obviously takes much longer." (F, middle manager) |
|  | Institutional support |  | **Normalisation of unsupported/unfunded activities/program**<br>"It is already considered part of the additional income allowance (TPP). From the Head of DHO to staff, no other incentives are allowed." (F, middle manager)<br>"Then, regarding infrastructure and equipment — to be honest, not every staff member here has their own PC or laptop. Only colleagues who have can use them independently. When there are technical problems, staff have to wait for others to finish their reporting tasks before they can use the available computer. Besides that, internet connectivity is honestly almost always a challenge for us in completing our work. Nowadays, nearly all reporting is application-based." (SY, middle manager)<br>"And fortunately, our colleagues are still willing to use their personal mobile data, | **Constraint in Human Resource planning**<br>"In my opinion, in issues like this [Human Resources] also require guidance and support, so that we can accurately calculate staffing needs. Because no matter how good the system we create is, if the workforce is limited, it will still be difficult to implement." (F, middle manager)<br>**Normalisation of unsupported/unfunded activities (within DHO)**<br>"...even without a specific budget, it can still run ... because it is integrated into each division." (C, top manager)<br>"there are no additional incentives or honorarium... that this is part of our job" (F, middle manager)<br>"It's not that this is perceived as a burden, but rather that the volume of work is simply too high. These tasks are actually part of their core responsibilities, but because the number of staff is limited, it ends up feeling heavy. In my view, this is a major challenge for the Health Office." (F, middle manager)<br>**Perceived asymmetrical reciprocity**<br>"There has been no cost-sharing support so far; the DHO has not received budgetary support from us. For example, when |
|  |  |  | so this has not become a major obstacle yet. However, it remains a challenge moving forward — how we can ensure that all staff are properly supported to access the internet reliably.” (SY, middle manager)<br>**Capacity strengthening to support data accountability**<br>“We need capacity strengthening ... so the data in the Linktree can be accountable and useful.” (SL, middle manager) | there were activities requiring shared costs, there was no such support. The DHO conveyed this concern to me. I raised it with the Jakarta office because it was beyond my authority. But HSI said it was not possible.” (HSI) |
|  | Perceived benefits, importance, relevance | **Expectations of benefits/costs (potential cost)** “If this adds more workload, it becomes a challenge for us.” (SL, middle manager)<br>**(data integration and connectivity )** “Hopefully with this one-data system, the data can be centralized, so it will make things easier.” (SL, middle manager) | **Adjustment cost**<br>“At the beginning, processing, inputting, and analysing data felt heavy because we had never done it before. It took a lot of time, but eventually it will make things easier.” (D, staff) | **Improved motivation and ownership**<br>“There has been a change in mindset ... managing and communicating data becomes easier ... even simple data can have a big impact.” (Y, staff) |
| Accountability | Power relations | **Learning-oriented accountability:**<br>“We were not pressured to be perfect; the focus was on learning.” (F, middle manager) | **Limited upward accountability:**<br>“The internalization process, including the SOP, is still lacking; honestly, I have only just seen it [the excel data bank format] once.” (I, middle manager) | **Leadership-dependent accountability:**<br>“...through the heads of divisions to remind ... so control is layered.” (F, middle manager)<br>“...if the Head of Office is not available ... there will be no routine meetings.” (SL, middle manager)<br>“...Others said, ‘We’re willing to carry this through to the analysis stage as expected, but please don’t give us additional tasks.’” (F, middle manager) |
|  | Norms and regulations | **Unsupportive practice/norms to the pursuit of accountability**<br>“It is hard to decide priorities if data are scattered.” (F, middle manager)<br>**Reliance on personal motivation**<br>“If the head of the Puskesmas is not proactively looking for the data, we may not aware of it. But in the Ministry of Health system, planning, implementation, and evaluation are all under core management, so we’re supposed to rely on that management unit.” (SL, middle manager) | **Data governance and standardization**<br>“My suggestion for the capacity building is to strengthen governance and data standardisation, whether they are discussions of the RPJMN or RPJMD and existing strategic plans.” (SL, middle manager)<br>**Norms of Performance Monitoring**<br>“There has already been a shared agreement. When we talk about performance, we also need to see the process. Maybe this month the data is not filled in, next month it is, and the third month it is not again, so the process becomes visible.” (C, top manager) | **Obligatory compliance**<br>“The deadline [to fill out the data format] is set to the 5th. We try to fill according to the indicators, as much as possible” (IA, middle manager)<br>“It has become our responsibility since it was officially launched and the decree was signed. Even if there is a revision, this is already running” (IA, middle manager) |
|  | Institutional support |  |  | “We have reached the end stage but the process is left behind, the foundation is still lacking” (F, middle manager)<br>**Workload constrained accountability**<br>“Some also think, ‘Even if I’m sick, no one will take care of my work.’ So they feel there’s no need to push themselves too hard....” (F, middle manager) |
|  | Perceived benefits, importance, relevance | **Data-informed planning & prioritization**<br>“It is hard to decide priorities if data is scattered.” (F, middle manager)<br>“We want the data to be used for planning, not just reporting.” (SL, middle manager) | **Objective Performance Measurement**<br>“In the coordination meeting, I said that everything now has to be measured objectively. This is one of the instruments to measure performance objectively.” (C, top manager) |  |

*Note: within the endline column, "Health Program and DHO" summarizes illustrative* *quotes describing how programme divisions and the DHO relate to each other* *operationally, while "Influence and strategic relationships" presents the research team's* *interpretive synthesis of the power and relationship dynamics those quotes illustrate.*

### Centralized hierarchy enabled rapid formal institutionalization but concentrated operational ownership

The timing of the Health Data Bank initiative coincided with the appointment of a new Head of the DHO, creating a favourable leadership environment for rapid institutional uptake.. Although externally initiated, the program was rapidly embraced by DHO leadership, particularly because it aligned with the Head of DHO’s longstanding interest in strengthening digital health information systems since his tenure as director of the district hospital. In interviews, the head of DHO consistently framed the initiative as part of a broader institutional effort to strengthen evidence-based decision-making and organizational innovation.

This strong leadership endorsement accelerated the initiative’s structural institutionalization. The Head of DHO routinely reinforced implementation during internal meetings, reminded divisions about reporting responsibilities, and positioned the initiative as a collective organizational commitment, not a temporary external project. By the endline phase, the initiative had acquired its own internal identity through the locally developed branding “MATA SIDIK,” reflecting growing symbolic legitimacy within the organization. As one division head explained, *“It has become our responsibility since it was officially launched and the decree was signed. Even if there is a revision, this is already running.”* (IDI_4).

However, while hierarchical leadership facilitated rapid formal endorsement, operational ownership remained concentrated within a small number of actors. Although responsibilities were formally distributed across five organizational units, implementation and monitoring functions became heavily centralized within the Secretariat. The secretary of the data bank management team emerged as the primary operational coordinator, responsible for monitoring compliance, following up with divisions, consolidating data, and communicating with the external technical team.

The effectiveness of this arrangement was reinforced by strong interpersonal alignment between the secretary and the Head of DHO. The secretary described their relationship as having “good chemistry” developed through years of working together in the district hospital, which facilitated communication and administrative coordination. At the same time, this concentration of operational authority limited broader organizational ownership. Divisions demonstrated varying levels of readiness and engagement, while some newly appointed department heads were still adapting to the initiative during the endline phase.

### Expanding analytical responsibilities without workload redistribution and adequate resources constrained operationalization

A recurring finding across all implementation phases was the tension between data bank responsibilities and existing workload demands within the DHO. Staff members were simultaneously responsible for multiple programs, administrative duties, and reporting obligations directed toward provincial and national authorities. The Health Data Bank was introduced as an additional layer of responsibility without corresponding redistribution of workload, staffing, or institutional resources.

Participants consistently described the density of existing reporting requirements and overlapping responsibilities across units. As one informant explained, “*We have to prepare annual reports, profiles, audits, many documents to complete.”* (IDI_1). Another participant highlighted the multi-program burden experienced at service delivery level, “*One Puskesmas staff member does not handle only one program.”* (IDI_5).

Leadership attempted to reduce resistance by strategically assigning personnel whose data bank responsibilities aligned with their existing job descriptions. Nevertheless, the initiative still required additional time for data consolidation, validation, communication, and analysis beyond routine administrative tasks. Staff frequently described implementation delays caused by competing urgent activities, including audit preparation, district leadership visits, and mandatory reporting deadlines from higher administrative levels.

No dedicated budget or additional personnel were allocated specifically for the initiative. Participation was explicitly framed as part of routine professional obligations, not compensated project work. As one participant stated, “*There are no additional incentives or honorarium… this is part of our job.”* (IDI_1)

While most participants did not reject the initiative itself, many expressed concern about the cumulative workload burden created by expanding responsibilities without corresponding institutional support. One participant explained, “*It’s not that this is perceived as a burden, but rather that the volume of work is simply too high. These tasks are actually part of their core responsibilities, but because the number of staff is limited, it ends up feeling heavy.”* (FGD)

### Perceived strategic value strengthened engagement and learning momentum despite incomplete implementation

Despite operational challenges and incomplete routine implementation, many participants expressed optimism regarding the long-term value of the Health Data Bank initiative. Across interviews and FGDs, staff frequently described the initiative as important for strengthening evidence-based planning, improving organizational coordination, and fostering a stronger culture of data use within the DHO. The initiative was not only understood as a technical reporting system, but increasingly perceived as part of a broader institutional transformation toward data-driven governance.

Leadership played an important role in shaping this perception by framing the initiative within a larger narrative of organizational innovation. Several participants referred to the implementation period as an ‘innovation year,’ reflecting a broader institutional push toward modernization and system improvement within the DHO. This framing contributed to a sense of collective purpose and encouraged staff to view participation in the data bank as part of organizational progress, not merely as an additional administrative task.

The initiative also appeared to stimulate gradual shifts in mindset regarding the importance of data analysis and utilization. Capacity-building activities exposed staff to concepts related to data communication, interpretation, and evidence-based decision-making, areas that had previously received less attention in routine program implementation. Participants increasingly recognized that data should not only be collected for reporting purposes but also analysed to identify problems and inform programmatic responses. As one participant noted, divisions had already begun conducting internal discussions and clarification processes regarding data quality and interpretation before escalating issues to the Secretariat.

Several participants also viewed the initiative as an opportunity to strengthen analytical capacity within the DHO. Even though routine operational systems were still developing, staff began to appreciate the potential strategic value of integrated data systems for planning, monitoring, and evaluation. Some expressed hope that once fully operational, the system would reduce fragmentation across programs and improve organizational responsiveness to health problems.

At the same time, perceived relevance was not distributed evenly across organizational units. Some participants questioned whether the primary benefits of the system would be experienced broadly across departments or remain concentrated at the leadership level. Discussions during interviews occasionally framed the initiative in terms of supporting managerial decision-making for the Head of DHO, particularly through statements emphasizing how integrated data would enable leadership to “*make decisions on health problems.*” While such views reflected recognition of the strategic importance of the system, they also suggested that some staff perceived the initiative as more directly serving upper-level managerial functions than routine operational needs at division or service-delivery levels.

This uneven perception of relevance influenced engagement across units. Divisions that more clearly recognized the usefulness of integrated data for their own programmatic responsibilities appeared more actively engaged in implementation, while others remained more compliance-oriented. Nevertheless, even among participants who acknowledged operational difficulties, there was broad recognition that strengthening data systems represented an important institutional direction for the DHO.

These findings suggest that perceived strategic value and organizational legitimacy helped sustain engagement during the early implementation period, even before routine operational benefits became fully visible. The initiative generated learning momentum and gradual shifts toward a stronger data culture despite incomplete institutionalization of everyday operational practices.

### Formal compliance was achieved, while operational activation was postponed

The initiative achieved structural institutionalization, but functional institutionalization remained incomplete. By the endline phase, the Health Data Bank had achieved a meaningful degree of structural formalization. The data team was constituted and maintained, SOPs had been formally endorsed and internally socialized, hardware had been procured, and capacity-building activities had been completed across all 25 team members, with substantial improvements in self-assessed and observed competencies in data communication and leadership.

By the endline phase, routine data submission, validation, and evaluation forum activities had not yet been fully implemented. The integrated database was preparing for operational launch, and divisions were beginning to engage with routine data submission cycles. The evaluation forum, designed to support review and action on data bank outputs, was scheduled for future activation. Quantitative indicators of data utilization were expected to improve as routine cycles of collection, consolidation, review, and discussion are established.

Participants recognized this gap and described it in terms of sequencing: analytical and communication capacities had been built before the foundational data processes that would give those capacities something to work with. As one staff member put it, “*We have reached the end stage but the process is left behind, the foundation is still lacking*” (FGD_Informant 3). Flexibility in SOP application, while pragmatically useful during the formative phase, also reduced accountability for deadline compliance: *“The deadline is set to the 5th. We try to fill according to the indicators, as much as possible*” (IDI_2). Governance structures, SOPs, teams, and training were established, yet routine operational behaviours had not become embedded into organizational practice.

### External partnership support facilitated implementation but also shaped expectations of reciprocity

HSI played an important role in initiating and legitimizing the Health Data Bank within the DHO. Participants described how the initiative emerged from HSI’s operational need for integrated health indicators, which initially required repeated communication with multiple focal points across divisions. The proposed “single-entry” data system was subsequently perceived as beneficial not only for HSI, but also for the DHO’s own internal data management and decision-making needs.

The external involvement of HSI also facilitated early implementation through technical assistance, coordination, and system development support. Participants generally described communication with HSI as open and collaborative, particularly through relationships built with senior district leadership and key coordinators within the DHO.

The findings also suggest that authority within the partnership extended beyond formal bureaucratic structures. HSI was widely perceived as possessing valuable technical expertise, external networks, and experience in health systems strengthening. This form of epistemic authority appeared to enhance the credibility of the initiative and facilitated acceptance among DHO actors. At the same time, the DHO retained formal decision-making authority over implementation. The partnership was not a simple donor-recipient relationship; it was characterized by interdependent forms of authority, in which bureaucratic legitimacy and technical expertise were mutually reinforcing. This dynamic may help explain both the rapid acceptance of the initiative and the expectations of reciprocity that emerged during implementation.

At the same time, the partnership also shaped expectations regarding reciprocity and institutional support. Some participants perceived that the DHO had invested substantial organizational effort in responding to repeated data requests and supporting implementation processes, while HSI’s institutional boundaries limited its ability to provide broader operational or financial support beyond formally designated activities. One participant described this not as an issue of “ethics,” but rather of “reciprocity,” reflecting expectations of more balanced institutional exchange within the partnership.

These findings suggest that externally initiated reforms may derive important legitimacy and implementation momentum from partner involvement, while also generating expectations regarding mutual support that extend beyond the technical scope of the intervention itself.

## Discussion

This study set out to examine why a health data bank initiative that attracted strong institutional endorsement, completed its capacity-building program, and established all the required governance instruments was able to achieve substantial structural institutionalization but had not yet transitioned into routine operational use within its initial implementation period. Importantly, the initiative achieved several intermediate outcomes, including formal governance arrangements, strengthened data management capacity, enhanced organizational ownership, and the establishment of a foundation for district performance management. The answer, we argue, lies not in technical inadequacy but in the political economy of the organizational context into which the reform was introduced [15,16]. Bureaucratic hierarchy accelerated formal compliance while concentrating motivational authority at the top; workload was intensified without redistribution; financial institutionalization was deferred; and the result was a system that achieved structural form without functional activation. Each of these dynamics is examined below in relation to the broader literature.

These findings extend Bossert’s decision space framework by demonstrating how decision space is shaped in practice by political economy dynamics, specifically bureaucratic hierarchy, workload distribution, and institutional incentives. While decision space is often conceptualized across authority, resources, and accountability, our analysis shows that misalignment between centralized authority and limited resource autonomy can produce structural institutionalization without functional operationalization.

The role of leadership in initiating and sustaining the Health Data Bank aligns with a well-established pattern in health system reform literature: top-down mandates can rapidly generate formal compliance, but they do not automatically produce the distributed ownership that sustains routine use [7,9]. In this case, the head of DHO’s demand for integrated decision-support was the primary catalyst for reform initiation, and his ongoing involvement sustained momentum across phases through routine reminders, monitoring expectations, and institutional framing. This is consistent with findings from district health system reforms in Ghana and Kenya, where strong district management leadership has been identified as a critical enabling factor for district-level data system reforms [19,20].

However, the same hierarchical dynamic that enabled rapid endorsement also created a structural vulnerability. When motivation and accountability are concentrated at the level of a single leader, reforms become contingent on that individual’s continued presence and attention. Participants’ recognition that routine meetings would cease without the head of DHO’s involvement is a signal not of staff disengagement but of a reform design that had not yet embedded accountability into horizontal processes or peer-level incentives. Bossert’s decision space framework is instructive here: authority was exercised at the top of the hierarchy but not meaningfully redistributed downward, limiting the functional autonomy of division-level coordinators to act independently [17]. The practical implication is that leadership endorsement, while necessary, must be deliberately accompanied by mechanisms that diffuse ownership, including clear role-based accountability, peer monitoring, and visible benefits at the division level. Although formal accountability mechanisms were established through the decree, SOPs, and team structure, these arrangements primarily supported structural institutionalization. They did not automatically generate routine demand for data, redistribute analytical responsibilities, or create incentives for sustained use, which are essential conditions for functional institutionalization.

The concentration of monitoring authority within the Secretariat reflected both organizational convention and a pragmatic response to the early-stage nature of the initiative. In the short term, this centralization provided coherence and enabled communication upward to leadership. However, it also limited the development of analytical ownership at the division level, the organizational stratum where data ultimately need to be prepared, interpreted, and used. This finding resonates with broader critiques of centralized HSI governance in LMICs, where single-point control over data consolidation tends to reproduce upward accountability flows instead of supporting horizontal learning and problem-solving [1,3].

Critically, while functionally effective in the short term, the reliance on a strong interpersonal relationship between the data bank secretary and the head of DHO represents a fragile coordination mechanism. Reforms that depend on specific dyadic relationships instead of institutionalized processes are inherently vulnerable to personnel change, a risk that is particularly acute in Indonesian district health offices where civil service rotation is common. As Bossert’s framework suggests, sustainable reform requires that authority be matched with commensurate accountability structures at each level, not concentrated at one node of a hierarchical system [21]. Moving toward distributed analytical ownership will require deliberate delegation of decision-making authority to division coordinators, supported by clear performance expectations and feedback mechanisms.

Perhaps the most consistent and politically significant finding in this study is the systematic externalization of reform costs onto staff labour. Data bank responsibilities were added to existing roles without any reduction in other duties, without dedicated resources, workload adjustment, or formal role restructuring, and without formal workload restructuring. This pattern is widely documented in health system reform literature and is theorized within street-level bureaucracy frameworks as a source of reform fatigue and partial implementation: when frontline workers absorb the costs of new systems without corresponding support, compliance becomes selective and intermittent [22,23].

In the Indonesian district context, this dynamic is compounded by the structural reality of multi-program staffing, where a single officer may carry simultaneous reporting obligations to national platforms across several vertical programs. The addition of a district-level data bank, however relevant, represents yet another layer in an already dense reporting environment. This is precisely what Wickremasinghe et al. [1] describe as the “layering” problem in LMIC health information systems: new initiatives are added without simplifying or consolidating existing demands, increasing burden instead of rationalizing effort. Without deliberate workload restructuring that includes the possibility of role specialization or dedicated data management positions, the sustainability of routine data bank operations will remain dependent on intrinsic motivation that, as participants themselves acknowledged, has limits.

The gap between structural formalization and functional activation observed in this study is not unique to West Sumbawa, but this case makes the distinction unusually visible. All the formal components of institutionalization were present by the endline: a constituted team, endorsed SOPs, a database structure, trained staff. Yet none of the functional indicators, such as routine data submission, evaluation forum activation, data-informed decision-making, had been achieved. This maps onto a distinction that implementation science has begun to articulate more precisely: the difference between adoption on paper and use in practice, or between what Fixsen et al. [24] term “paper implementation” and “full implementation.” In addition, the initiative fostered district ownership through the Mata Sidik identity, strengthened data-related competencies among staff, and established governance structures intended to support future performance management processes.

From a political economy perspective, this gap is explained not by capacity deficits but by the absence of institutional conditions that make routine use the path of least resistance. When data submission competes with urgent reporting deadlines and the operational benefits of routine use have not yet been realized, it will consistently lose. While participants recognized the potential benefits of the Health Data Bank, including improved access to integrated indicators and support for district-level decision-making, these benefits had not yet translated into observable operational gains through routine use. When the evaluation forum that should create demand for data bank outputs has never been activated, the system has no internal market for its own products. This creates a self-reinforcing equilibrium: without routine use, benefits remain invisible; without visible benefits, motivation to sustain routine use remains weak. Breaking this equilibrium requires deliberate activation. Not of structural components, but of the behavioural and organizational routines that give those components a function. Similar dynamics have been documented in district HIS reforms in sub-Saharan Africa and South Asia, where dashboard functionality preceded behavioural change by months or years, and where the transition to routine use required specific facilitation beyond technical deployment [6,25,26].

In this sense, the findings reinforce a broader distinction that is often underemphasized in health information system reforms: institutionalization should not be equated with the presence of formal structures, but with the embedding of new routines into organizational practice. Structural compliance, manifested through team establishment, SOP endorsement, and system availability, may satisfy external accountability requirements, but does not in itself guarantee functional use. Without corresponding shifts in authority distribution, workload organization, and incentive structures, reforms risk remaining operationally dormant despite appearing institutionally complete. This distinction is critical for both evaluators and implementers, as it reframes success from the establishment of systems to their sustained use in everyday decision-making.

The findings provide several considerations for policymakers, district managers, and development partners seeking to strengthen district-level data systems and performance management processes. Most immediately, reform implementers and development partners should resist the assumption that structural compliance, the establishment of teams, SOPs, and databases, constitutes successful institutionalization. Monitoring frameworks should be redesigned to measure behavioural activation alongside structural readiness, tracking whether routine submission cycles are operating, whether evaluation forums are generating documented action points, and whether data outputs are visibly informing planning decisions. These behavioural indicators are harder to observe than structural ones, but they are the only meaningful evidence that a reform has become part of organizational life. Job or workload analysis must be undertaken before formally assigning the tasks to certain

More fundamentally, reforms that introduce new data responsibilities into district health offices must grapple seriously with the political economy of workload. Adding analytical functions to already burdened civil servants without dedicated resources, role specialization, or explicit workload relief is a design choice with predictable consequences. Development partners and district governments should explore mechanisms for financial institutionalization, including dedicated budget lines for data management within annual district planning documents, as well as structural options such as designated data officer roles that reduce the fragmentation of responsibilities across multi-program staff. The lesson from West Sumbawa is not that the Health Data Bank was poorly conceived, but that its institutional conditions were insufficiently prepared. Technical readiness and human capacity are necessary but not sufficient; what is equally required is deliberate redistribution of authority, workload, and resources across the organizational hierarchy.

Redistribution of authority and workload requires a combination of formal and procedural adjustments, including the delegation of analytical decision-making to division-level actors, the establishment of routine data review forums that foster horizontal accountability, and the alignment of data responsibilities with existing workloads through role designation and reporting integration. Without such measures, data reforms risk reinforcing existing hierarchical and workload imbalances instead of supporting the routine use of data for district performance management and decision-making.

Our findings also suggest that externally supported reforms may derive early legitimacy and momentum from partner involvement. However, sustaining implementation beyond the initial phase requires a transition to locally embedded ownership and accountability. This aligns with broader evidence that the sustainability of health system reforms depends not only on technical support, but also on the integration of new practices into routine institutional processes [27,28].

This study has several limitations. First, as a single-district case study, the findings may not be fully generalizable to other settings with different institutional arrangements, although they provide transferable insights for similar decentralized health systems. Second, data were collected as part of a program evaluation in close collaboration with the District Health Office, which may have introduced social desirability bias, with participants potentially emphasizing positive aspects of the initiative or moderating criticism. Third, the longitudinal design captured implementation over a relatively short period of about five months, focusing on early institutionalization phases. Comparable data system reforms typically show visible operational benefits only after several cycles of routine use, often a year or more, so this study may not fully reflect the longer-term dynamics of routine use and sustainability that a longer observation period would reveal. Fourth, while multiple data sources were used, the analysis relied on qualitative interpretation, which is inherently shaped by researchers’ perspectives. To enhance analytical rigor, coding and interpretation were conducted by multiple researchers, with regular discussions to refine code definitions and resolve discrepancies. Triangulation across interviews, focus group discussions, observations, and document review was used to strengthen the credibility of findings and reduce reliance on any single data source. Nevertheless, some degree of interpretive bias may remain. Finally, as the analysis represents a secondary interpretation of evaluation data through a political economy lens, there is a risk that certain themes were emphasized over others. This was addressed through systematic coding, use of a predefined conceptual framework, and transparent documentation of analytical decisions. However, alternative interpretations of the data are possible.

## Conclusion

This study shows that the institutionalization of district-level health data systems is not solely a technical process, but an organizational one shaped by authority, workload, and incentives. In West Sumbawa, the Health Data Bank achieved structural formalization, through established teams, governance instruments, and capacity building, but had not yet transitioned into routine operational use. This gap reflects not a lack of technical readiness, but the absence of distributed ownership, workload alignment, and dedicated financing.

The findings suggest that hierarchical systems can facilitate reform endorsement, but may limit broader ownership and routine activation if not accompanied by mechanisms that distribute responsibility and sustain engagement. Within performance management cycle, the Health Data Bank has functioned as a structural backbone, but has not yet been fully activated as a routine decision-making tool. For district health system strengthening, particularly in the context of ILP implementation, these findings highlight the need to move beyond structural readiness toward functional use. Embedding data systems into routine practice requires aligning decision-making authority with resources, integrating new tasks into existing workflows, and securing long-term institutional support.

## Data Availability

The datasets generated and/or analyzed during the current study are not publicly available due to ethical restrictions and the need to protect participant confidentiality. Anonymized data are available from the corresponding author upon reasonable request and subject to ethical approval requirements.

## Acknowledgment

The authors would like to thank the Health Data Bank Team of the West Sumbawa District Health Office for their valuable support and collaboration throughout the study. The authors also thank Health Systems Insight (HSI) for its technical support in initiating and implementing the Health Data Bank. We also acknowledge the contributions of all participants who generously shared their experiences and perspectives during the evaluation process. During the preparation of this work, the firs author used Claude (Anthropic) to improve the language and readability of the manuscript. After using this tool, the author(s) reviewed and edited the content as needed and take full responsibility for the content of the published article.

## Notes

### Competing Interest Statement

The authors have declared no competing interest.

### Author Declarations

The study received ethics approval from the Internal Review Board of the Faculty of Medicine, Public Health, and Nursing at Universitas Gadjah Mada, Indonesia (Approval Number: KE/FK/1275/EC 2025).

## References

1. Wickremasinghe D, Hashmi IE, Schellenberg J, Avan BI. District decision-making for health in low-income settings: a systematic literature review. Health Policy Plan. 2016;31(suppl_2):ii12–ii24.

2. Roman TE, Cleary S, McIntyre D. Exploring the Functioning of Decision Space: A Review of the Available Health Systems Literature. International Journal of Health Policy and Management. 2017;6(7):365–376. doi:10.15171/ijhpm.2017.26.

3. Bishai D, Paina L, Li Q, Peters DH, Hyder AA. Advancing the application of systems thinking in health: why cure crowds out prevention. Health Research Policy and Systems. 2014;12:28. doi:10.1186/1478-4505-12-28.

4. Dehnavieh R, Haghdoost A, Khosravi A, Hoseinabadi F, Rahimi H, Poursheikhali A, et al. The District Health Information System (DHIS2): a literature review and meta-synthesis of its strengths and operational challenges based on the experiences of 11 countries. Health Inf Manag. 2019;48(2):62–75.

5. Odei-Lartey EO, Prah RKD, Anane EA, Danwonno H, Gyaase S, Oppong FB, et al. Utilization of the national cluster of district health information system for health service decision-making at the district, sub-district and community levels in selected districts of the Brong Ahafo region in Ghana. BMC Health Serv Res. 2020;20(1):514. doi:10.1186/s12913-020-05349-5.

6. Gimbel S, Baynes C, Tilahun Alemu H, Hirschhorn L, et al. Barriers and facilitators to data use for decision making: the experience of the African Health Initiative partnerships in Ethiopia, Ghana, and Mozambique. Glob Health Sci Pract. 2022;10(Suppl 1):e2100666. doi:10.9745/GHSP-D-21-00666.

7. Sheikh K, Witter S, Schleiff M. Learning health systems in low-income and middle-income countries: exploring evidence and expert insights. BMJ Global Health. 2022;7(Suppl 7):e008115. doi:10.1136/bmjgh-2021-008115.

8. Karuveettil V, Janakiram C, Ramesh S, Ramachandran A, Mathur M, Varma B, et al. Political economy analysis of health: a scoping review of concepts, definitions, frameworks, outcomes, and applications. Health Policy Plan. 2026;41(Suppl 1):i91–i110. doi:10.1093/heapol/czaf096.

9. Ndlovu J, Mbandlwa Z. Institutional dynamics of health system strengthening in low- and middle-income countries: A review of governance mechanisms and reform challenges. Societies. 2026;16(6):176. doi:10.3390/soc16060176.

10. Kigume R, Maluka S, Kamuzora P. Health sector decentralisation in Tanzania: How do institutional capacities influence use of decision space? Int J Health Plann Manage. 2018;33(4):e1050–e1066. doi:10.1002/hpm.2587.

11. Bulthuis SE, Kok MC, Raven J, Dieleman MA. Factors influencing the scale-up of public health interventions in low- and middle-income countries: A qualitative systematic literature review. Health Policy and Planning. 2020;35(2):219–234.

12. Health Systems Insight. Case study: integrated primary health care reform. Jakarta: Health Systems Insight; 2025. Report No.: HSIPUB-0062.

13. Carter N, Bryant-Lukosius D, DiCenso A, Blythe J, Neville AJ. The use of triangulation in qualitative research. Oncology nursing forum. 2014;41(5):545–547. doi:10.1188/14.ONF.545-547.

14. Fritz V, Kaiser K, Levy B. Problem-driven governance and political economy analysis: good practice framework. Washington, DC: World Bank; 2009.

15. Kuwawenaruwa A, Mollel H, Machonchoryo JM, Margini F, Jaribu J, Binyaruka P. A political economy analysis of strengthening health information system in Tanzania. BMC Medical Informatics and Decision Making. 2023;23(1):245. doi:10.1186/s12911-023-02319-9.

16. Bossert TJ, Mitchell AD. Health sector decentralization and local decision-making: decision space, institutional capacities and accountability in Pakistan. Soc Sci Med. 2011;72(1):39–48. doi:10.1016/j.socscimed.2010.10.019.

17. Bossert T. Analyzing the decentralization of health systems in developing countries: decision space, innovation, and performance. Soc Sci Med. 1998;47(10):1513–1527. doi:10.1016/s0277-9536(98)00234-2.

18. Braun V, Clarke V. One size fits all? What counts as quality practice in (reflexive) thematic analysis? Qual Res Psychol. 2021;18(3):328–352. doi:10.1080/14780887.2020.1769238.

19. Heerdegen ACS, Aikins M, Amon S, Agyemang SA, Wyss K. Managerial capacity among district health managers and its association with district performance: A comparative descriptive study of six districts in the Eastern Region of Ghana. PLoS ONE. 2020;15(1):e0227974. doi:10.1371/journal.pone.0227974.

20. Oware PM, Omondi G, Adipo C, Adow M, Wanyama C, Odallo D, et al. Perceived accuracy and utilisation of DHIS2 data for health decision making and advocacy in Kenya: A qualitative study. PLOS Global Public Health. 2025;5(8):e0004508. doi:10.1371/journal.pgph.0004508.

21. Bossert T, Beauvais JC. Decentralization of health systems in Ghana, Zambia, Uganda, and the Philippines: a comparative analysis of decision space. Health Policy Plan. 2002;17(1):14–31. doi:10.1093/heapol/17.1.14.

22. Lipsky M. Street-level bureaucracy: dilemmas of the individual in public services. New York: Russell Sage Foundation; 1980.

23. Singh V, Chandwani R, Pingali V, Dalal A. Frontline workers in India’s tuberculosis (TB) elimination efforts: a street-level bureaucracy perspective. BMC Public Health. 2025;25(1):4412. doi:10.1186/s12889-025-25431-z.

24. Fixsen DI, Naoom SF, Blasé KA, Friedman RM, Wallace F. Implementation research: a synthesis of the literature. Tampa, FL: University of South Florida, Louis de la Parte Florida Mental Health Institute, The National Implementation Research Network; 2005. (FMHI Publication #231).

25. Delamou A, Grovogui FM, Camara F, Kolié D, Goumou T, Miller L, et al. Implementation of the national community health policy in Guinea: a decision space analysis of the roles and responsibilities of community health workers. Bundesgesundheitsblatt - Gesundheitsforschung - Gesundheitsschutz. 2025;68(7):738–746. doi:10.1007/s00103-025-04076-8.

26. Schellenberg J, Marchant T, Persson LÅ, Avan BI, Dubale M, Taye G. Data-driven decision-making for district health management: a cluster-randomised study in 24 districts of Ethiopia. BMJ Global Health. 2024;9(2):e014140. doi:10.1136/bmjgh-2023-014140.

27. Scheirer MA, Dearing JW. An agenda for research on the sustainability of public health programs. American Journal of Public Health. 2011;101(11):2059–2067.

28. Sarriot E, Winch PJ, Ryan LJ, Edison J, Bowie J, Swedberg E, et al. A methodological approach and framework for sustainability assessment in NGO-implemented primary health care programs. International Journal of Health Planning and Management. 2004;19(1):23–41.

